# Clinical Outcomes, Management, Healthcare Resource Utilization, and Cost According to the CHA_2_DS_2_-VASc Scores in Asian Patients with Nonvalvular Atrial Fibrillation

**DOI:** 10.1101/2024.04.10.24305638

**Authors:** Keye Fan, Yue Xiao, Aoming Xue, Jifang Zhou

## Abstract

**Background:** The prognosis among non-valvular atrial fibrillation (NVAF) patients with different CHA_2_DS_2_-VASc scores in the contemporary Asian population remains unclear. Additionally, there is a lack of research examining the disparities in management patterns, healthcare resource utilization (HCRU), and cost among these patients. Therefore, this study aims to assess the incidence of clinical outcomes in NVAF patients with different CHA2DS2-VASc scores and explore their management patterns, HCRU, and cost.

**Methods and Results:** This retrospective cohort study assessed patients diagnosed with NVAF between January 2018 and July 2022, utilizing a merged dataset from China. Patients were stratified into 3 cohorts by CHA_2_DS_2_-VASc scores: low-risk (0 for males, 1 for females), intermediate-risk (1 for males, 2 for females), and high-risk (≥2 for males, ≥3 for females). One-year incidence rates of clinical outcomes (including ischemic stroke, transient ischemic attack, arterial embolism, and major bleeding) were calculated as events per 100 person-years. Cumulative incidence and crude and adjusted hazard ratios (aHRs) with 95% confidence intervals (CIs) were calculated using the Fine and Gray models. Management patterns, HCRU, and cost were analyzed descriptively. Among 419,490 NVAF patients (mean age: 75.2 years, 45.1% female), 16,541 (3.9%) were classified as low-risk, 38,494 (9.2%) as intermediate-risk, and 364,455 (86.9%) as high-risk. The mean (SD) age-adjusted Charlson comorbidity index score was 4.7 (2.0), increasing with CHA_2_DS_2_-VASc scores. The one-year cumulative incidence of ischemic stroke was 3.2% (95% CI, 2.9%–3.5%) for low-risk, 4.9% (95% CI, 4.7%–5.2%) for intermediate-risk (aHR, 1.3, 95% CI, 1.2–1.4), and 12.2% (95% CI, 12.1%–12.3%) for high-risk (aHR, 2.5, 95% CI, 2.3–2.8). Meanwhile, the incidence of transient ischemic attack, arterial embolism, and major bleeding showed a similar increasing trend from low-risk to high-risk. Within one year after the index date, 16.4% of patients in the low-risk cohort received oral anticoagulants (OACs), while 11.1% of patients in the high-risk cohort received OACs. The mean (SD) number of all-cause hospitalizations was 0.1 (0.1), 0.1 (0.2), and 0.1 (0.3) per-patient-per-month (PPPM) for low-risk, intermediate-risk, and high-risk, respectively. The mean (SD) length of stay increased from 0.9 (1.1) days PPPM for the low-risk to 1.2 (1.8) days PPPM for the high-risk.

**Conclusion:** This study demonstrates that contemporary Asian NVAF patients with higher CHA_2_DS_2_-VASc scores experience higher incidence of adverse outcomes and increased hospital resource consumption. There is insufficient utilization of OACs and other AF management measures across all CHA_2_DS_2_-VASc scores groups. These findings provide new evidence for improving patient management and guiding resource allocation in healthcare.

**Clinical Perspective:** *What Is New?:*

- This large-scale study assessed the incidence of adverse clinical outcomes among contemporary Asian atrial fibrillation patients by CHA_2_DS_2_-VASc scores.
- This study demonstrated suboptimal management across all CHA_2_DS_2_-VASc score groups, with higher hospital resource utilization observed in groups with higher CHA_2_DS_2_-VASc scores.

*What Are the Clinical Implications?:*

- This study indicated that AF patients with elevated CHA_2_DS_2_-VASc scores face a greater risk of adverse clinical outcomes.
- Our findings informed decision-making on healthcare resource allocation and AF management.

## Introduction

Atrial fibrillation (AF) is the most common sustained cardiac arrhythmia worldwide. The global prevalence of AF was reported at 4,977 cases per million people in 2017, and it was higher in men than in women.^1^ In China, the estimated prevalence of AF ranged from 0.69% to 2.31% and increased with age.^2–4^ As a chronic cardiovascular disease, AF is associated with other conditions, including hypertension, diabetes, myocardial infarction, congestive heart failure, coronary artery disease, and sleep apnea.^5,6^ Meanwhile, AF is associated with a spectrum of adverse clinical outcomes, most notably a fivefold escalation in the risk of stroke, imposing an escalating disease burden.^7,8^ Effective anticoagulation management, particularly the use of oral anticoagulants (OACs), has been shown to significantly reduce the risk of stroke in patients with AF.^9^

The CHA_2_DS_2_-VASc score, including congestive heart failure, hypertension, age 75 years or older, diabetes, stroke, vascular disease, age 65 to 74 years, and sex category, is a commonly used tool for assessing the risk of stroke in patients with AF. This scoring system has been recommended by guidelines for preventing stroke and is considered a crucial factor in deciding whether to prescribe OACs to individuals with AF.^7,10^ Previous studies have shown a higher incidence of adverse clinical outcomes among nonvalvular atrial fibrillation (NVAF) patients with higher levels of CHA_2_DS_2_-VASc risk scores.^11–16^ However, these studies were primarily based on Western populations or relied on limited sample sizes or data from years past. Considering the differences in ethnicity and the evolving spectrum of diseases, the incidence of adverse outcomes among contemporary Asian NVAF patients with different CHA_2_DS_2_-VASc scores remains unclear.^17,18^Additionally, research suggests that Asian populations experience suboptimal AF management and exhibit greater disparities in healthcare burden between AF and non-AF individuals.^19,20^ However, to our knowledge, there is currently no study that granulates the differences in management patterns, healthcare resource utilization (HCRU), and cost by CHA_2_DS_2_-VASc scores, which could potentially offer insights for medical decision-making and resource allocation.

Therefore, the main objective of this study is to assess the incidence of major clinical outcomes in contemporary Asian NVAF patients with different CHA_2_DS_2_-VASc scores using a combined dataset, as well as to describe the treatment patterns, HCRU, and cost among these patients.

## Method

### Data source

The dataset utilized in this study was extracted from the Jiangsu Population Health Records Big Data Platform in China. Established in 2012 by the Provincial Health Commission, this information platform encompasses 218 tertiary hospitals province-wide, offering comprehensive medical and health data, including inpatient and outpatient records, prescription records, and surgical records, primarily coded using the International Classification of Diseases, 9th Revision (ICD-9) or 10^th^ Revision (ICD-10). Additionally, unstructured medical orders and mortality information are also integrated. Data availability determined the study period from 2018 to 2022.

### Study design

This study employed a retrospective cohort design and adhered to the Strengthening the Reporting of Observational Studies in Epidemiology (STROBE) reporting guidelines. The cohort of AF cases was identified by having at least one inpatient admission or two outpatient visits with a gap exceeding 30 days with a diagnosis of AF during the study period (from January 1, 2018 to July 31, 2022). AF was defined by the diagnosis code (ICD-9, 427.31; ICD-10, I48), or medical terms in Chinese. The AF index date was defined as the earliest date of diagnosis during the study period (the diagnosis date was defined as the discharge date for hospital discharge records and the outpatient date for outpatient records). The duration preceding the AF index date was defined as the baseline period. Patients who had invalid demographic information or were aged < 18 years at the AF index date were excluded. Patients who had a valvular AF diagnosis from hospital discharge records, outpatient records, or surgery records during the baseline period were also excluded.

NVAF patients were further categorized into 3 cohorts by CHA_2_DS_2_-VASc risk scores: low-risk (0 for males, 1 for females), intermediate-risk (1 for males, 2 for females), and high-risk (≥2 for males, ≥3 for females), which were captured and assessed from hospital discharge records and outpatient records using the ICD-10 codes or medical terms during the baseline period (Table S1).

### Baseline patient characteristics

Patient demographic characteristics, including age, gender, and year of index date, were obtained at the index date. Common comorbidities, including coronary artery disease, heart failure, myocardial infarction, atherosclerosis, cerebrovascular disease, hypertension, diabetes, dyslipidemias, chronic pulmonary diseases, chronic liver disease, renal disease, pneumonia, malignancy, autoimmune disease, anemia, depression, dementia, frailty, and sleep apnea during the baseline period, were extracted from hospital discharge records and outpatient records for each patient using the ICD-10 codes or medical terms (Table S2). Both the Charlson comorbidity index (CCI) scores and the age-adjusted Charlson comorbidity index (ACCI) scores (plus 1 point for every decade aged 50 and over, up to a maximum of 4 points) were calculated (Table S3).

### Follow-up clinical outcomes

The follow-up period for each patient ranged from the AF index date up to one year, the date of death, or the end of the study (July 31, 2022), whichever came first. The observation period was limited to a maximum of one year due to the dynamic nature of stroke risk factors.^21^ We investigated four main clinical outcomes: ischemic stroke (IS), transient ischemic attack (TIA), arterial embolism (AE), and major bleeding (MB). All outcomes were identified from hospital discharge records and outpatient records using the ICD-10 codes or medical terms (Table S4).

### Patient management, HCRU, and cost

Medication information (including warfarin, rivaroxaban, dabigatran, apixaban, and edoxaban) was captured from prescription records using the generic name of each drug. Patients with an AF index date after May 31, 2022, were excluded from the analysis of medication usage due to the unavailability of prescription records.

Nonpharmacological strategies (including left atrial appendage closure [LAAC], catheter ablation, surgical ablation, and pacemaker implantation) relevant to AF management were captured from surgical records using the ICD-9 codes or medical terms (Table S5).

HCRU included outpatient visits, inpatient admissions, and inpatient length of stay (LOS). The cost was the total cost of one hospitalization and was converted to 2022 US dollars from the Chinese yuan (CNY). All HCRU and cost were evaluated from the hospital discharge records or outpatient records and presented as per patient per month (PPPM).

### Statistical analysis

Patient baseline characteristics, management, HCRU, and cost were analyzed descriptively. Means (SD) or medians (IQR) were calculated for continuous variables. Counts and frequencies were calculated for categorical variables. A Kruskal-Wallis test was used as appropriate for group comparisons. The one-year incidence rates for different outcomes were determined by computing the number of events per 100 person-years (PYs) with 95% confidence intervals (CIs). The cumulative incidences of clinical outcomes were computed utilizing the Fine and Gray model, which accounted for death as a competing risk. Additionally, to conduct further comparisons among cohorts, hazard ratios (HRs) with corresponding 95% CIs were calculated employing both a crude model and an adjusted model, adjusting for age, sex, year of diagnosis, and CCI scores. A two-sided P-value of <0.05 was considered statistically significant. Analyses were performed using R version 4.2.1.

### Supplementary analysis

In light of the lower age threshold for increased stroke risk among Asian AF patients, the latest Chinese AF management guidelines have proposed the CHA_2_DS_2_-VASc-60 stroke risk scoring system.^22^ Within this framework, patients aged 60 to 64 years are assigned 1 point, while those aged 65 years or older receive 2 points. Therefore, we conducted a supplementary analysis to assess the incidence of clinical outcomes among NVAF patients stratified by CHA_2_DS_2_-VASc-60 scores.

## Results

### Baseline characteristics

A total of 419,490 NVAF patients were included in the final study cohort. According to the CHA_2_DS_2_-VASc scores, 16,541 (3.9%) individuals were classified as low-risk, 38,494 (9.2%) as intermediate-risk, and 364,455 (86.9%) as high-risk (Figure 1). The baseline characteristics of the patients were presented in Table 1. The mean (SD) age of the overall population was 75.2 (11.0) years. The proportion of female patients increased with the CHA_2_DS_2_-VASc risk scores, ranging from 37.0% in the low-risk group to 46.3% in the high-risk group. Hypertension was the most prevalent comorbidity, followed by heart failure, atherosclerosis, and cerebrovascular disease, accounting for 57.1%, 46.2%, 37.1%, and 36.3%, respectively. The mean (SD) CCI scores and ACCI scores were 1.7 (1.5) and 4.7 (2.0), respectively, both increasing with the CHA_2_DS_2_-VASc scores.

**Figure 1.**
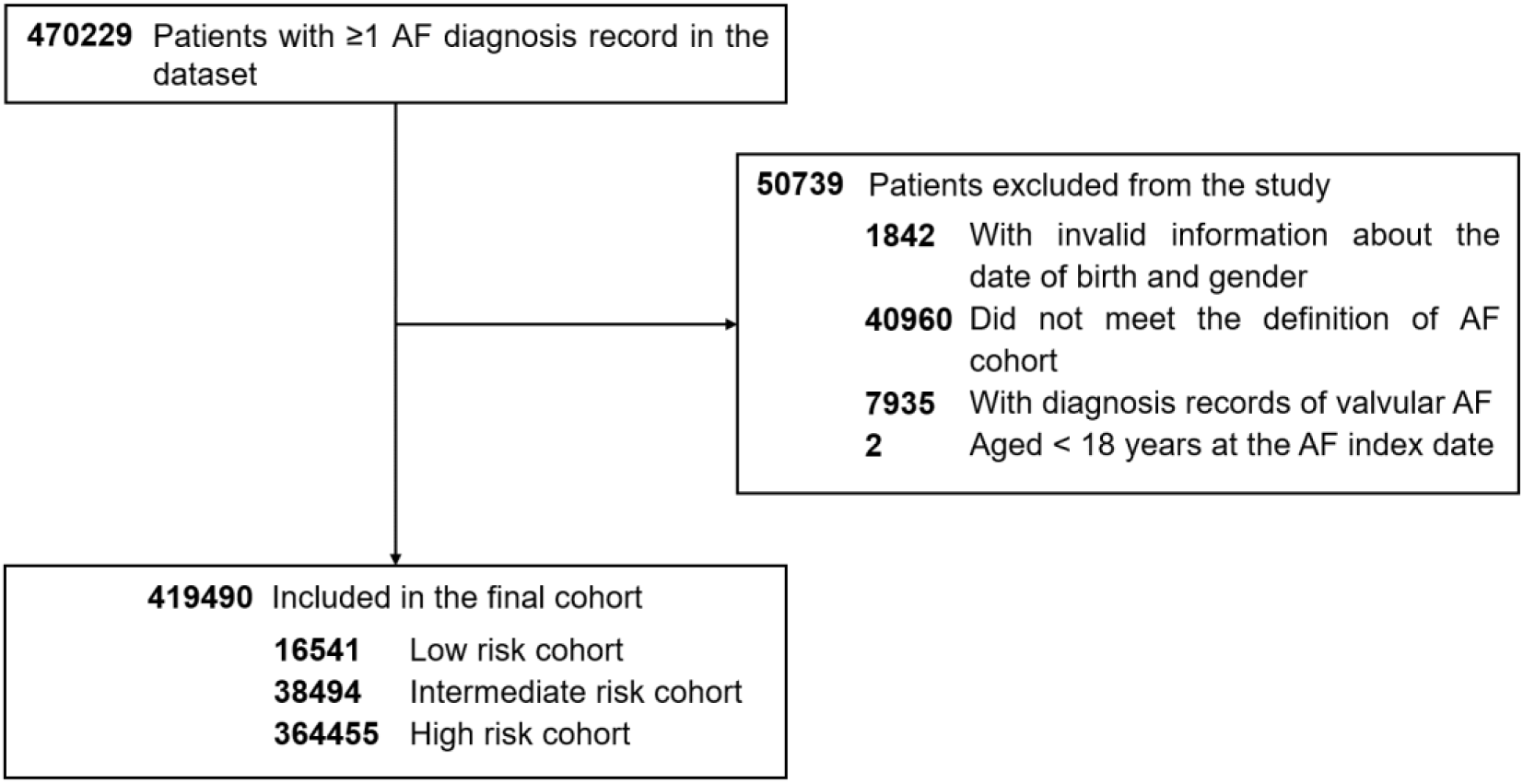
Flow chart of study cohort. Abbreviations: AF, atrial fibrillation.

**Table 1.**
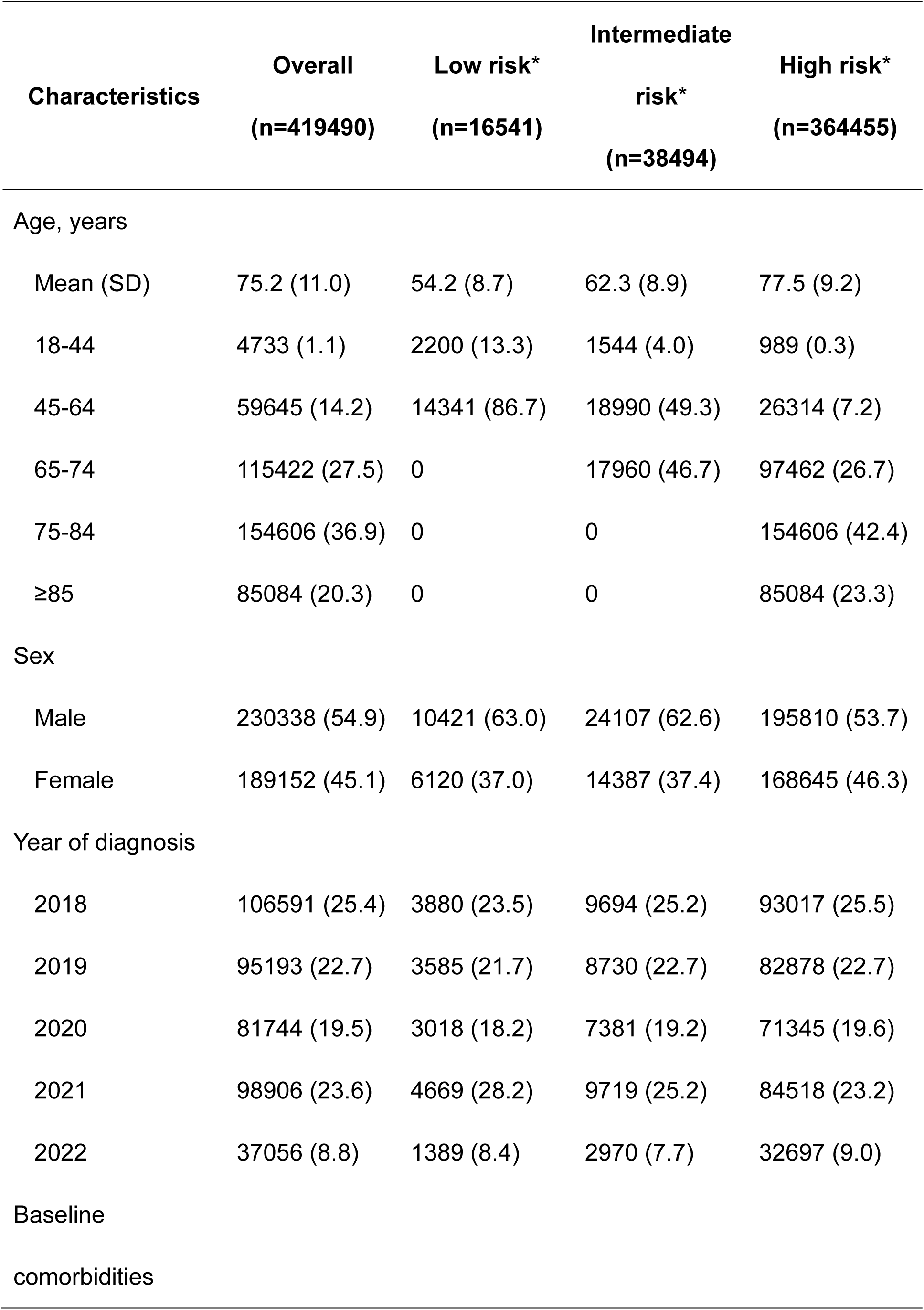
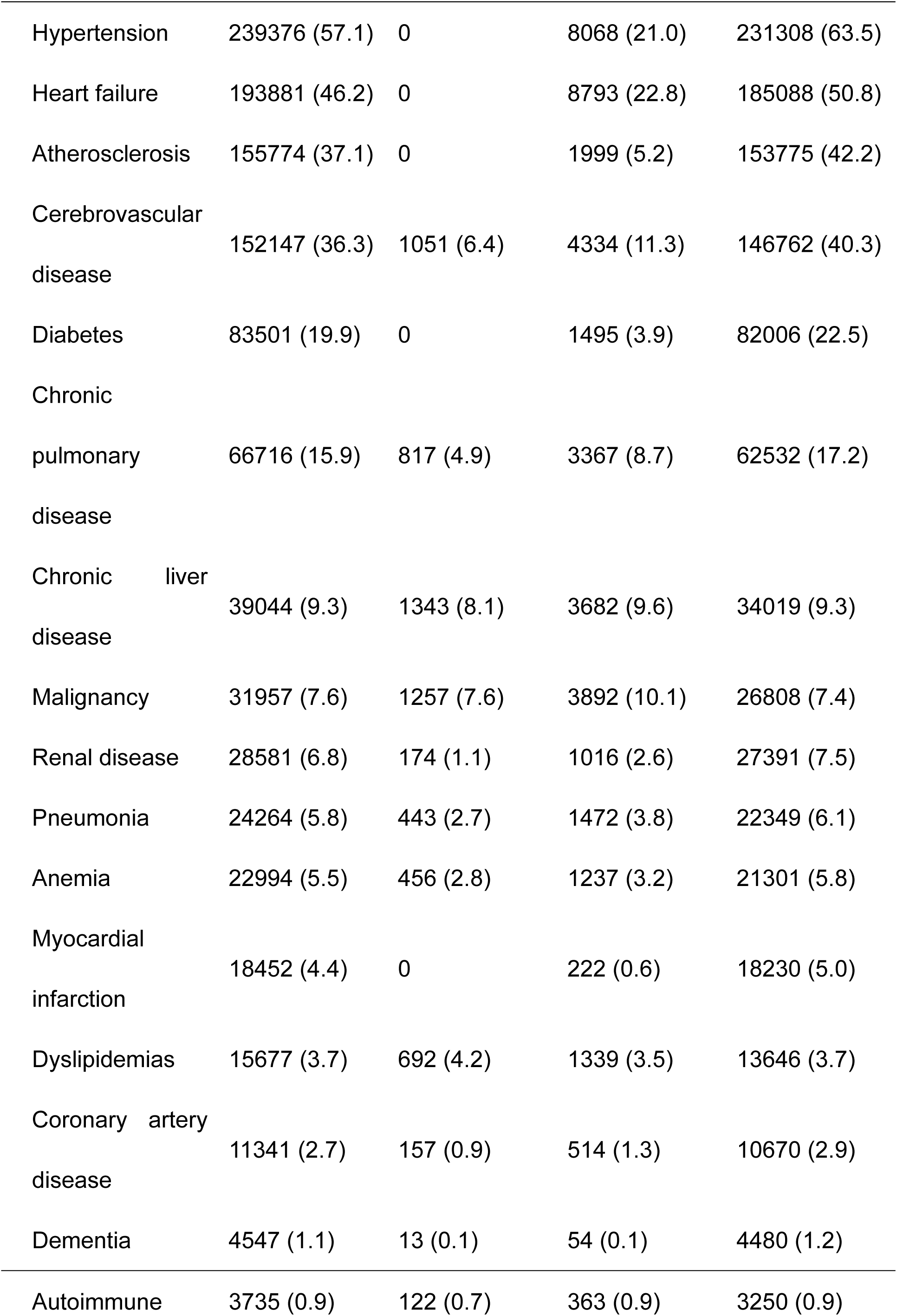
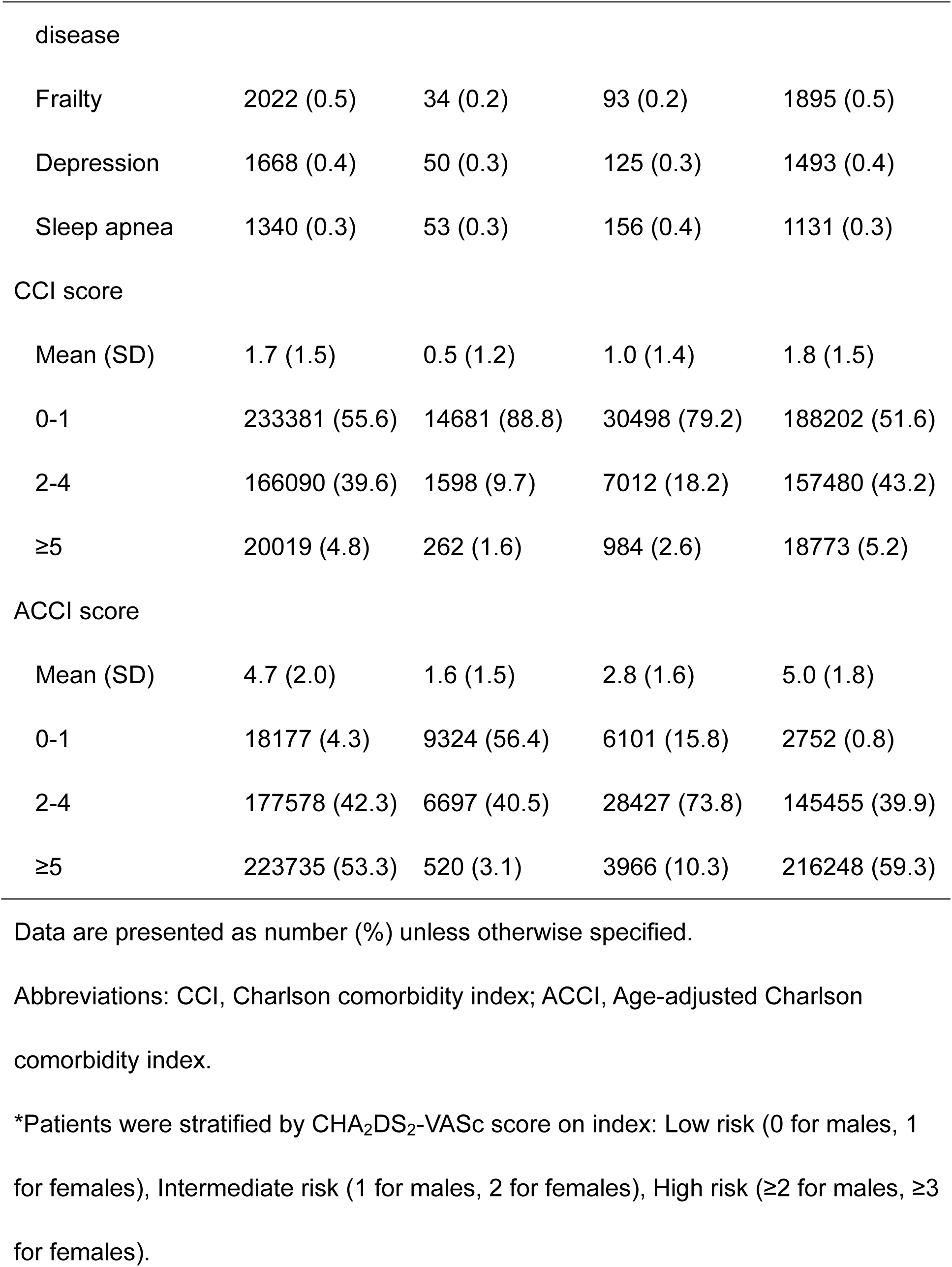
Baseline characteristics of atrial fibrillation patients.

| **Characteristics** | **Overall**<br>**(n=419490)** | **Low risk\***<br>**(n=16541)** | **Intermediate**<br>**risk\***<br>**(n=38494)** | **High risk\***<br>**(n=364455)** |
| --- | --- | --- | --- | --- |
| Age, years |  |  |  |  |
| Mean (SD) | 75.2 (11.0) | 54.2 (8.7) | 62.3 (8.9) | 77.5 (9.2) |
| 18-44 | 4733 (1.1) | 2200 (13.3) | 1544 (4.0) | 989 (0.3) |
| 45-64 | 59645 (14.2) | 14341 (86.7) | 18990 (49.3) | 26314 (7.2) |
| 65-74 | 115422 (27.5) | 0 | 17960 (46.7) | 97462 (26.7) |
| 75-84 | 154606 (36.9) | 0 | 0 | 154606 (42.4) |
| ≥85 | 85084 (20.3) | 0 | 0 | 85084 (23.3) |
| Sex |  |  |  |  |
| Male | 230338 (54.9) | 10421 (63.0) | 24107 (62.6) | 195810 (53.7) |
| Female | 189152 (45.1) | 6120 (37.0) | 14387 (37.4) | 168645 (46.3) |
| Year of diagnosis |  |  |  |  |
| 2018 | 106591 (25.4) | 3880 (23.5) | 9694 (25.2) | 93017 (25.5) |
| 2019 | 95193 (22.7) | 3585 (21.7) | 8730 (22.7) | 82878 (22.7) |
| 2020 | 81744 (19.5) | 3018 (18.2) | 7381 (19.2) | 71345 (19.6) |
| 2021 | 98906 (23.6) | 4669 (28.2) | 9719 (25.2) | 84518 (23.2) |
| 2022 | 37056 (8.8) | 1389 (8.4) | 2970 (7.7) | 32697 (9.0) |
| Baseline comorbidities |  |  |  |  |
| Hypertension | 239376 (57.1) | 0 | 8068 (21.0) | 231308 (63.5) |
| Heart failure | 193881 (46.2) | 0 | 8793 (22.8) | 185088 (50.8) |
| Atherosclerosis | 155774 (37.1) | 0 | 1999 (5.2) | 153775 (42.2) |
| Cerebrovascular<br>disease | 152147 (36.3) | 1051 (6.4) | 4334 (11.3) | 146762 (40.3) |
| Diabetes | 83501 (19.9) | 0 | 1495 (3.9) | 82006 (22.5) |
| Chronic<br>pulmonary<br>disease | 66716 (15.9) | 817 (4.9) | 3367 (8.7) | 62532 (17.2) |
| Chronic liver<br>disease | 39044 (9.3) | 1343 (8.1) | 3682 (9.6) | 34019 (9.3) |
| Malignancy | 31957 (7.6) | 1257 (7.6) | 3892 (10.1) | 26808 (7.4) |
| Renal disease | 28581 (6.8) | 174 (1.1) | 1016 (2.6) | 27391 (7.5) |
| Pneumonia | 24264 (5.8) | 443 (2.7) | 1472 (3.8) | 22349 (6.1) |
| Anemia | 22994 (5.5) | 456 (2.8) | 1237 (3.2) | 21301 (5.8) |
| Myocardial<br>infarction | 18452 (4.4) | 0 | 222 (0.6) | 18230 (5.0) |
| Dyslipidemias | 15677 (3.7) | 692 (4.2) | 1339 (3.5) | 13646 (3.7) |
| Coronary artery<br>disease | 11341 (2.7) | 157 (0.9) | 514 (1.3) | 10670 (2.9) |
| Dementia | 4547 (1.1) | 13 (0.1) | 54 (0.1) | 4480 (1.2) |
| Autoimmune | 3735 (0.9) | 122 (0.7) | 363 (0.9) | 3250 (0.9) |
| disease |  |  |  |  |
| Frailty | 2022 (0.5) | 34 (0.2) | 93 (0.2) | 1895 (0.5) |
| Depression | 1668 (0.4) | 50 (0.3) | 125 (0.3) | 1493 (0.4) |
| Sleep apnea | 1340 (0.3) | 53 (0.3) | 156 (0.4) | 1131 (0.3) |
| CCI score |  |  |  |  |
| Mean (SD) | 1.7 (1.5) | 0.5 (1.2) | 1.0 (1.4) | 1.8 (1.5) |
| 0-1 | 233381 (55.6) | 14681 (88.8) | 30498 (79.2) | 188202 (51.6) |
| 2-4 | 166090 (39.6) | 1598 (9.7) | 7012 (18.2) | 157480 (43.2) |
| ≥5 | 20019 (4.8) | 262 (1.6) | 984 (2.6) | 18773 (5.2) |
| ACCI score |  |  |  |  |
| Mean (SD) | 4.7 (2.0) | 1.6 (1.5) | 2.8 (1.6) | 5.0 (1.8) |
| 0-1 | 18177 (4.3) | 9324 (56.4) | 6101 (15.8) | 2752 (0.8) |
| 2-4 | 177578 (42.3) | 6697 (40.5) | 28427 (73.8) | 145455 (39.9) |
| ≥5 | 223735 (53.3) | 520 (3.1) | 3966 (10.3) | 216248 (59.3) |

Data are presented as number (%) unless otherwise specified.
Abbreviations: CCI, Charlson comorbidity index; ACCI, Age-adjusted Charlson comorbidity index.
\*Patients were stratified by CHA<sub>2</sub>DS<sub>2</sub>-VASc score on index: Low risk (0 for males, 1 for females), Intermediate risk (1 for males, 2 for females), High risk (≥2 for males, ≥3 for females).

### Clinical outcomes within one year after AF index date

The one-year incidence rates for IS, TIA, AE, and MB among the overall population were 12.4 (95% CI, 12.3–12.5), 1.1 (95% CI, 1.0–1.1), 0.5 (95% CI, 0.5–0.5), and 3.1 per 100 PYs (95% CI, 3.1–3.2), respectively. Meanwhile, the incidence rates for the four investigated clinical outcomes showed an increasing trend from the low-risk group to the high-risk group, namely from 3.4 (95% CI, 3.1–3.7) to 13.7 per 100 PYs (95% CI, 13.5–13.8) for IS, from 0.5 (95% CI, 0.4–0.6) to 1.1 per 100 PYs (95% CI, 1.1–1.2) for TIA, from 0.2 (95% CI, 0.1–0.3) to 0.5 per 100 PYs (95% CI, 0.5–0.6) for AE, and from 1.6 (95% CI, 1.4–1.8) to 3.3 per 100 PYs (95% CI, 3.2–3.3) for MB (Table 2).

**Table 2.** Prognosis within one year after the diagnosis of atrial fibrillation.

| **Risk** | **Events** | **PYs** | **Incidence rate (95% CI),**<br>**per 100 PYs** | **Cumulative incidence**<br>**(95% CI), %** | **Crude HR**<br>**(95% CI)** | **Adjusted HR\***<br>**(95% CI)** |
| --- | --- | --- | --- | --- | --- | --- |
| Ischemic stroke |  |  |  |  |  |  |
| Overall | 44576 | 358807 | 12.4 (12.3-12.5) | 11.2 (11.1-11.3) | - | - |
| Low risk | 506 | 14925 | 3.4 (3.1-3.7) | 3.2 (2.9-3.5) | 1 [Reference] | 1 [Reference] |
| Intermediate risk | 1803 | 34621 | 5.2 (5.0-5.5) | 4.9 (4.7-5.2) | 1.5 (1.4-1.7) | 1.3 (1.2-1.4) |
| High risk | 42267 | 309261 | 13.7 (13.5-13.8) | 12.2 (12.1-12.3) | 4.0 (3.6-4.3) | 2.5 (2.3-2.8) |
| Transient ischemic<br>attack |  |  |  |  |  |  |
| Overall | 4060 | 383127 | 1.1 (1.0-1.1) | 1.0 (1.0-1.1) | - | - |
| Low risk | 76 | 15188 | 0.5 (0.4-0.6) | 0.5 (0.4-0.6) | 1 [Reference] | 1 [Reference] |
| Intermediate risk | 191 | 35571 | 0.5 (0.5-0.6) | 0.5 (0.5-0.6) | 1.1 (0.8-1.4) | 1.0 (0.8-1.3) |
| High risk | 3793 | 332368 | 1.1 (1.1-1.2) | 1.1 (1.1-1.2) | 2.3 (1.8-2.8) | 2.0 (1.5-2.5) |
| Arterial embolism |  |  |  |  |  |  |
| Overall | 1874 | 384129 | 0.5 (0.5-0.5) | 0.5 (0.4-0.5) | - | - |
| Low risk | 32 | 15208 | 0.2 (0.1-0.3) | 0.2 (0.1-0.3) | 1 [Reference] | 1 [Reference] |
| Intermediate risk | 98 | 35602 | 0.3 (0.2-0.3) | 0.3 (0.2-0.3) | 1.3 (0.9-2.0) | 1.3 (0.9-2.0) |
| High risk | 1744 | 333319 | 0.5 (0.5-0.6) | 0.5 (0.5-0.5) | 2.5 (1.8-3.5) | 2.4 (1.6-3.6) |
| Major bleeding |  |  |  |  |  |  |
| Overall | 11802 | 378545 | 3.1 (3.1-3.2) | 3.0 (2.9-3.0) | - | - |
| Low risk | 243 | 15095 | 1.6 (1.4-1.8) | 1.6 (1.4-1.8) | 1 [Reference] | 1 [Reference] |
| Intermediate risk | 838 | 35177 | 2.4 (2.2-2.6) | 2.3 (2.1-2.4) | 1.5 (1.3-1.7) | 1.3 (1.1-1.5) |
| High risk | 10721 | 328273 | 3.3 (3.2-3.3) | 3.1 (3.0-3.2) | 2.0 (1.8-2.3) | 1.3 (1.1-1.5) |

Abbreviations: PY, person-year; HR, hazard ratio.
\*Adjusted for age, sex, year of diagnosis, and Charlson comorbidity index score.

The one-year cumulative incidence of all four clinical outcomes exhibited a trend similar to their respective incidence rates. (Table 2, Figure 2). When further comparing the differences among different CHA_2_DS_2_-VASc risk groups, except for the non-significantly higher incidence of TIA (aHR, 1.0, 95% CI, 0.8–1.3) and AE (aHR, 1.3, 95% CI, 0.9–2.0) in the intermediate-risk group compared to the low-risk group, all other results indicated a significant association between higher CHA_2_DS_2_-VASc scores and an increased incidence of clinical outcomes (Table 2).

**Figure 2.**
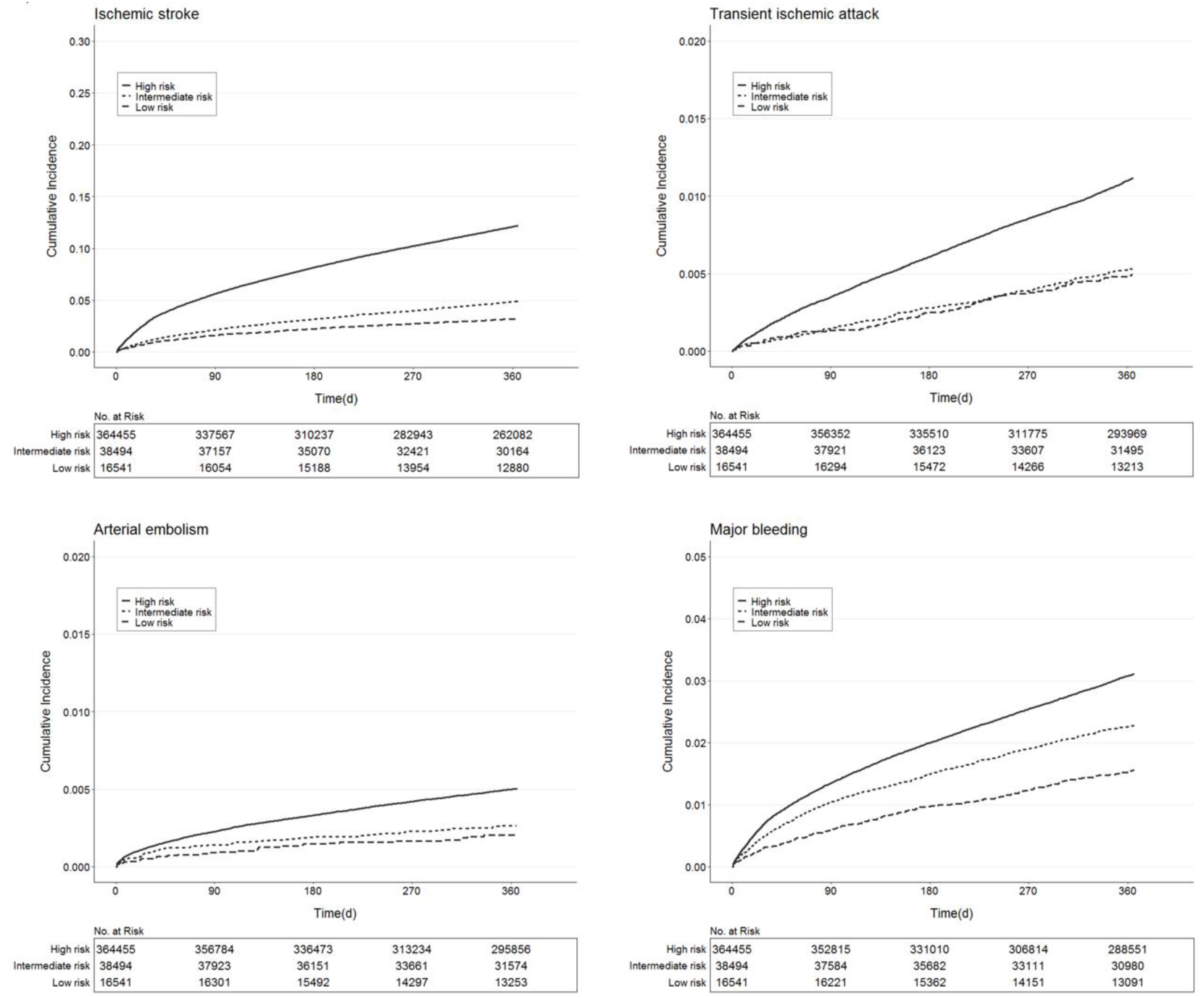
Cumulative incidence of clinical outcomes.

### Supplementary analysis

The estimated incidence of clinical outcomes among various risk groups based on the CHA_2_DS_2_-VASc-60 scores closely approximated those based on the CHA_2_DS_2_-VASc scores (Table 2, Table S6). However, the risk of AE in the intermediate-risk group was significantly higher than that in the low-risk group (aHR = 2.4, 95% CI, 1.3–4.4) based on the CHA_2_DS_2_-VASc-60 scores (Table S6).

### Patient management within one year after AF index date

In total, 11.7% of patients were prescribed OACs, and 8.6% were prescribed non-vitamin K oral anticoagulants (NOACs, namely rivaroxaban, dabigatran, apixaban, and edoxaban). The most commonly used OAC in AF patients was rivaroxaban, with a prescription rate of 6.6% during the follow-up period (Figure 3, Table S7). The group with higher CHA_2_DS_2_-VASc scores demonstrated a lower prescription rate of OACs, from 16.4% for the low-risk group to 11.1% for the high-risk group. In addition, in terms of nonpharmacological strategies, 1.4% of patients underwent catheter ablation, with a higher proportion in the low-risk group compared to the high-risk group (5.7% vs. 0.9%), and 0.4% underwent pacemaker implantation (Figure 3, Table S7).

**Figure 3.**
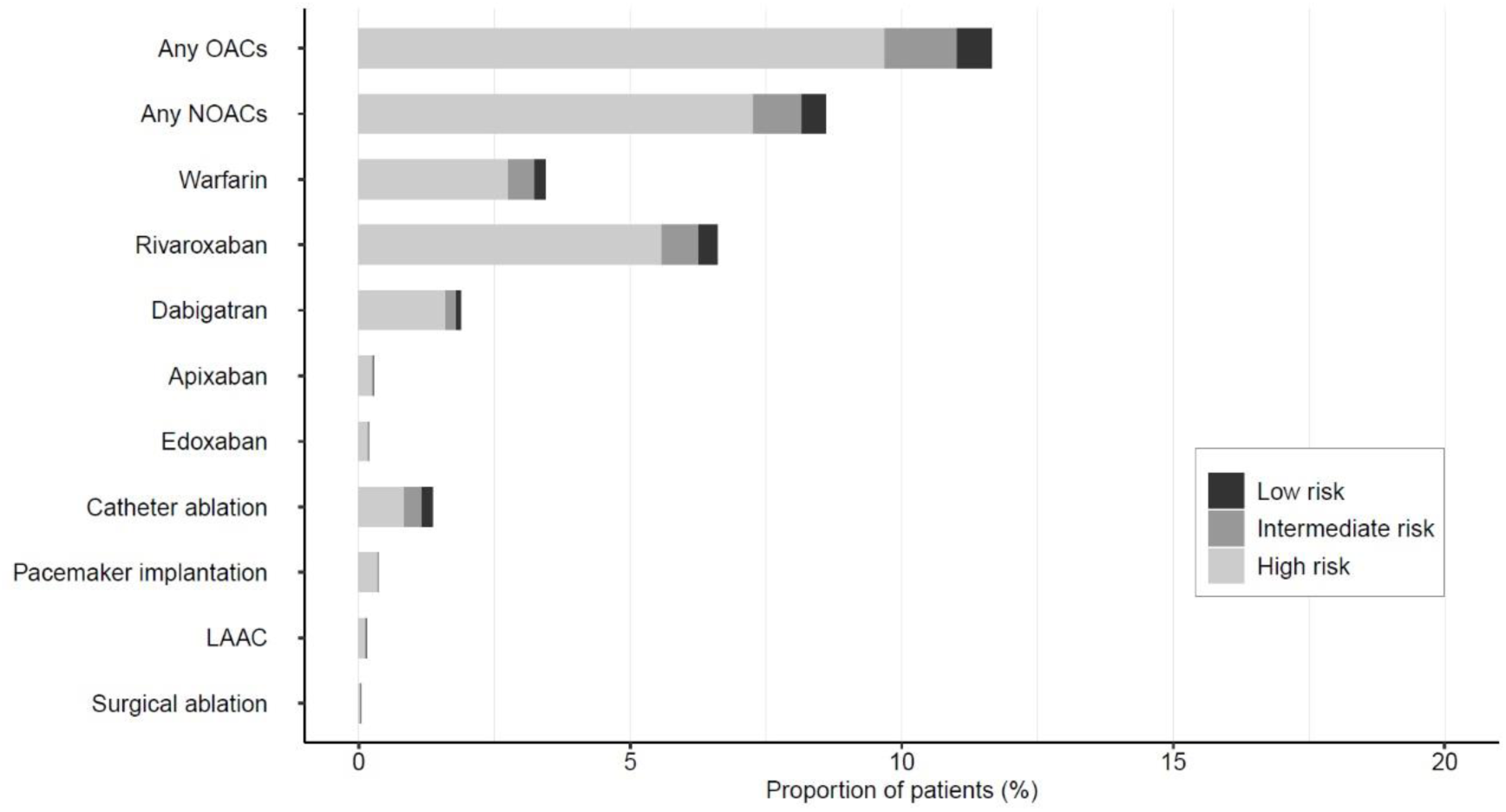
Management pattern of atrial fibrillation patients. Abbreviations: OAC, oral anticoagulant; NOAC, non-vitamin K oral anticoagulant; LAAC, left atrial appendage closure.

### HCRU and cost within one year after AF index date

Table 3 illustrates a comprehensive overview of HCRU and cost, both aggregated and stratified by CHA_2_DS_2_-VASc scores. The mean (SD) number of all-cause hospitalizations and all-cause outpatient visits were 0.1 (0.3) PPPM and 0.2 (0.5) PPPM, respectively. The mean (SD) LOS was 1.1 (1.7) days PPPM, and an increasing trend was observed with higher CHA_2_DS_2_-VASc scores, from 0.9 (1.1) days PPPM for the low-risk group to 1.2 (1.8) days PPPM for the high-risk group. The mean (SD) healthcare cost in the overall study population was $341.9 ($1630.7) PPPM, with $454.2 ($783.6) for the low-risk cohort, $383.9 ($1218.7) for the intermediate-risk cohort, and $334.2 ($1683.8) for the high-risk cohort.

**Table 3.** Health care resource utilization and cost stratified by CHA_2_DS_2_-VASc score.

|  | Overall<br>(n = 419490) | Low risk<br>(n = 16541) | Intermediate risk<br>(n = 38494) | High risk<br>(n = 364455) | p value* |
| --- | --- | --- | --- | --- | --- |
| Hospitalization |  |  |  |  |  |
| Number of patients<br>hospitalized, N (%) | 150779 (35.9) | 4475 (27.1) | 12386 (32.2) | 133918 (36.7) | - |
| All cause hospitalizations,<br>N | 316258 | 9340 | 27385 | 279533 | - |
| Mean (SD)<sup>†</sup> | 0.1 (0.3) | 0.1 (0.1) | 0.1 (0.2) | 0.1 (0.3) | < 0.001 |
| Median (IQR)<sup>†</sup> | 0.0 (0.0-0.1) | 0.0 (0.0-0.1) | 0.0 (0.0-0.1) | 0.0 (0.0-0.1) |  |
| Length of stay |  |  |  |  |  |
| Mean (SD)<sup>†‡</sup>, days | 1.1 (1.7) | 0.9 (1.1) | 1.0 (1.3) | 1.2 (1.8) | < 0.001 |
| Median (IQR)<sup>†</sup> | 0.8 (0.6-1.2) | 0.7 (0.5-1.1) | 0.7 (0.5-1.1) | 0.8 (0.6-1.2) |  |
| Cost of Hospitalization |  |  |  |  |  |
| Mean (SD)<sup>†‡</sup>, \$ | 341.9 (1630.7) | 454.2 (783.6) | 383.9 (1218.7) | 334.2 (1683.8) | < 0.001 |
| Median (IQR)<sup>†</sup> | 144.5 (88.2-288.1) | 170.9 (83.8-591.1) | 145.9 (83.9-352.5) | 143.8 (88.7-280.1) |  |
| Outpatient visits |  |  |  |  |  |
| Number of patients with an outpatient visit, N (%) | 119912 (28.6) | 6587 (39.8) | 13410 (34.8) | 99915 (27.4) | - |
| All cause outpatient visits, N | 745458 | 37976 | 83492 | 623990 | - |
| Mean (SD)<sup>†</sup> | 0.2 (0.5) | 0.2 (0.6) | 0.2 (0.5) | 0.2 (0.5) | < 0.001 |
| Median (IQR)<sup>†</sup> | 0.0 (0.0-0.1) | 0.0 (0.0-0.2) | 0.0 (0.0-0.2) | 0.0 (0.0-0.1) |  |

\**P* values calculated using a Kruskal-Wallis test.
<sup>†</sup>Results were calculated as per patient per month (PPPM).
<sup>‡</sup>Only patients with medical records within one year after atrial fibrillation diagnosis were included in the calculation.

## Discussion

This study was a large-scale retrospective cohort analysis encompassing a total of 419,490 NVAF patients recorded using the regional Population Health Big Data Platform. This study demonstrated an increased incidence of adverse clinical outcomes in contemporary Asian NVAF patients with higher CHA_2_DS_2_-VASc scores. Meanwhile, we observed that the proportion of patients who had utilized medications or nonpharmacological strategies for AF management was remarkably low. Furthermore, we investigated the HCRU and cost among NVAF patients stratified by CHA_2_DS_2_-VASc scores and found that patients in the high-risk group had higher inpatient resource consumption. The findings of this study emphasized the importance of focusing on NVAF patients with higher CHA_2_DS_2_-VASc scores and informed decision-making on healthcare resource allocation and AF management. The majority of patients in the study exhibited high CHA_2_DS_2_-VASc scores, with 86.9% of individuals being classified as high-risk. This result was consistent with a previous study based on five large health claims databases in the United States, where 89.0% of patients were estimated to have a high-risk of stroke.^12^Similar demographic characteristics, including an average age of over 70 years old and a higher proportion of males, were observed in comparable investigations.^14,23^

This study revealed a higher incidence of adverse clinical outcomes compared to previous studies and an increasing trend with higher CHA_2_DS_2_-VASc scores. The incidence of IS in Asian patients included in this study was notably higher across different CHA_2_DS_2_-VASc score categories compared to Western populations.^11,12^ Additionally, the estimated incidence of IS in our study was higher than those reported in most prior studies involving Asian populations, highlighting a greater disease burden for the Chinese population.^13–16^ We observed that the incidence of TIA and AE also increased with the CHA_2_DS_2_-VASc scores. However, there was no significant difference in the risk of TIA and AE between the intermediate-risk and low-risk groups, suggesting a limited utility of CHA_2_DS_2_-VASc scores in predicting the risk of TIA and AE in Asian patients with lower scores. Moreover, consistent with a study from Japan, a significant increase in the incidence of MB was observed from the low-risk group to the high-risk group.^13^

Given that Asian AF patients have a lower stroke age threshold, modified versions of the CHA_2_DS_2_-VASc score have been proposed. For instance, Taiwan introduced the MCHA_2_DS_2_-VASc score, which has been incorporated into international guidelines and considered a potentially advantageous alternative.^10,24^ In the latest Chinese AF management guidelines, the CHA_2_DS_2_-VASc-60 score was proposed for assessing stroke risk in Chinese AF patients.^22^ Therefore, we explored the clinical outcomes of AF patients with different CHA_2_DS_2_-VASc-60 scores in the supplementary analysis. The results revealed that the incidence among patients stratified by CHA_2_DS_2_-VASc-60 scores was comparable to those stratified by CHA_2_DS_2_-VASc scores. Notably, compared to the CHA_2_DS_2_-VASc scores, the intermediate-risk (aHR, 1.4 vs. 1.3) and high-risk groups (aHR, 3.0 vs. 2.5) stratified by the CHA_2_DS_2_-VASc-60 scores exhibited higher risks of IS. Additionally, the intermediate-risk group based on the CHA_2_DS_2_-VASc-60 scores showed a significantly higher risk of AE compared to the low-risk group (aHR, 2.4, 95% CI: 1.3– 4.4). Therefore, the CHA_2_DS_2_-VASc-60 scores may serve as a more suitable stroke risk assessment tool for the Chinese population, although further validation is required.

The findings of this study indicate suboptimal management of Asian NVAF patients across all CHA_2_DS_2_-VASc score strata. This study revealed a significant underutilization of OACs among NVAF patients. Throughout the follow-up period, only approximately 12% of patients received prescriptions for any OACs, and those who received prescriptions for any NOACs accounted for merely around 9%. The low utilization rate of OACs is widespread in China.^20,25^ This is primarily attributed to doctors’ concerns about the bleeding risk associated with OACs as well as suboptimal medication adherence among patients. Another reason for the low prescription rate may be the exclusion of data from retail pharmacies or non-tertiary hospitals, which may also partially account for the lower OAC utilization observed in the high-risk group compared to the low-risk group. Certain patients might acquire medications directly from retail pharmacies or non-tertiary hospitals, resulting in their prescription data being absent from this study’s records. In addition to assessing the use of OACs, this study examined other management strategies. The proportion of patients undergoing LAAC remained far less than 1%, with comparable results observed across all three cohorts. Notably, this study found a significantly higher proportion of patients in the low-risk group undergoing catheter ablation compared to the high-risk group (5.7% vs. 0.9%), indicating that younger patients are more likely to opt for catheter ablation for rhythm control. This is in line with the current understanding of rhythm control, namely that early implementation of catheter ablation leads to greater clinical benefits for patients.^10,26^

Moreover, this study offers the latest estimates of HCRU and cost for modern Asian NVAF patients, both collectively and categorized by CHA_2_DS_2_-VASc scores, potentially providing valuable evidence for healthcare resource allocation decisions. Approximately one-third of Asian patients included in this study experienced at least one hospitalization within one year after diagnosis, a proportion similar to that of the European (37.2%) and North American (37.5%) populations.^27^ Furthermore, our study indicated that patients with higher CHA_2_DS_2_-VASc scores have higher hospitalization resource needs. Compared to the low-risk group, patients in the high-risk group had higher hospitalization rates and longer LOS. Conversely, patients in the high-risk group exhibited lower outpatient visit rates compared to those in the low-risk group. This might be due to the increased consultation costs and reduced accessibility of tertiary hospitals. Consequently, older patients more likely classified into the high-risk group may prefer seeking outpatient care for common illnesses at primary healthcare facilities not covered in this study. Additionally, this study revealed an average inpatient cost of $341.9 PPPM, which is lower than observed in Western populations.^28,29^ This disparity is largely attributed to differences in socioeconomic determinants of health, such as financial resources, social support, and access to healthcare among different racial groups.^30^ Meanwhile, we noted that the average inpatient costs for patients in the low-risk group appeared to be higher than those in the high-risk group. A plausible explanation is the smaller sample size in the low-risk group, resulting in a more skewed distribution of cost (IQR for low-risk group, $83.8–$591.1 vs. high-risk group, $88.7–$280.1).

## Limitations

The study encountered several limitations that warrant acknowledgment. Firstly, its retrospective nature, relying on administrative databases not primarily intended for research, poses inherent constraints. The reliance on coded variables raises the possibility of coding errors and disease misclassifications, potentially introducing selection bias. Secondly, the absence of certain detailed patient information, such as electrocardiograms and specific subtypes of atrial fibrillation, could not be accessed, which might have provided valuable insights into the study outcomes. Thirdly, the study’s scope was confined to AF patients seeking treatment at tertiary medical institutions, limiting the generalizability of the findings to broader populations. Additionally, unmeasured confounders may have influenced the observed associations, as factors not accounted for in the study could potentially impact the outcomes under investigation. Furthermore, limited follow-up durations may have restricted the ability to capture long-term effects or changes in patient outcomes over time, potentially affecting the comprehensiveness of the study’s findings.

## Conclusions

Utilizing a dataset comprising more than 400,000 NVAF individuals, we conducted a comprehensive analysis of adverse clinical outcomes, management patterns, HCRU, and cost among contemporary Asian NVAF patients overall and stratified by CHA_2_DS_2_-VASc scores. Our study found that individuals with higher CHA_2_DS_2_-VASc scores had a higher incidence of adverse clinical outcomes and increased hospital resource consumption. Moreover, inadequate anticoagulation therapy and other management strategies were observed across all CHA_2_DS_2_-VASc score groups. With the exacerbation of population aging, an increase in the proportion of AF patients with higher CHA_2_DS_2_-VASc scores is foreseeable. Therefore, to alleviate the burden on both patients and society, more effective AF management strategies and the appropriate allocation of healthcare resources are warranted.

## Data Availability

The data are not publicly available due to the nature of administrative database.

## Non-standard Abbreviations and Acronyms

AF: Atrial fibrillation
OAC: Oral anticoagulant
NVAF: Nonvalvular atrial fibrillation
HCRU: Healthcare resource utilization
ICD: International Classification of Diseases
CCI: Charlson comorbidity index
ACCI: Age-adjusted Charlson comorbidity index
IS: Ischemic stroke
TIA: Transient ischemic attack
AE: Arterial embolism
MB: Major bleeding
LAAC: Left atrial appendage closure
LOS: Length of stay
PPPM: Per patient per month
PY: Person-year
CI: Confidence interval
HR: Hazard ratio
NOAC: Non-vitamin K oral anticoagulant

## Acknowledgments

We thank our colleagues for providing consolidated help in accomplishing this study. All authors participated in the study’s design. AX and YX undertook data cleaning duties. KF and JZ were responsible for the analysis of the data, the interpretation of the results, and the drafting of the manuscript.

## Sources of Funding

This research did not receive any specific grants from funding agencies in the public, commercial, or not-for-profit sectors.

## Disclosures

None.

## Supplemental Material

Tables S1–S7

## Notes

### Competing Interest Statement

The authors have declared no competing interest.

### Author Declarations

The study protocol received ethical approval from the Nanjing First Hospital Institutional Review Board (IRB) in Jiangsu, China (KY20230615-01-KS-01)

